# Quantifying morphological changes in middle trapezius with ultrasound scanning and a novel histogram-matching algorithm for adults with and without Facioscapulohumeral dystrophy (FSHD)

**DOI:** 10.1101/2024.01.11.24301162

**Authors:** Fraser Philp, Erik Meilak, Martin Seyres, Tracey Willis, Naomi Winn, Anand Pandyan

**Author notes:** Corresponding author Name: Dr Fraser Philp, Mailing address: School of Health Sciences, Thompson Yates Building, University of Liverpool, The Quadrangle, Brownlow Hill, Liverpool, L69 3GB. United Kingdom.

## Abstract

**Background and Objectives:** Facioscapulohumeral dystrophy (FSHD) is a neuromuscular disease causing changes in muscle structure that can negatively affect upper-limb function. Echogenicity, measured using quantified muscle ultrasound, is a potential biomarker that could be used for informing decision making. Histogram-matching allows for image normalisation, which could enable comparison of echogenicity between different machine capture settings which is a current limitation. This study aimed to investigate if ultrasonography and histogram-matching can measure trapezius muscle echogenicity and morphology for differentiating between people with and without FSHD, and different levels of arm function.

**Methods:** Single measurement timepoint case control study of adults with FSHD and age- and sex-matched controls. Main outcomes were trapezius echogenicity and muscle thickness measured using 2D-ultrasound, and maximum thoracohumeral elevation angle, measured using 3D-movement analysis. A sensitivity analysis evaluating the effect of histogram-matching and different reference images was conducted. Between group differences for echogenicity were evaluated using an unpaired student t-test. Echogenicity, muscle thickness and range of movement were plotted to evaluate the explained variance between variables.

**Results:** Data was collected for 14 participants (10M:4F), seven with FSHD and seven controls with a mean (SD) age of 41.6 (15.7). Normalisation was necessary and echogenicity values for the FSHD group were higher than the controls (118.2 (34.0) vs 42.3 (14.0) respectively, with statistically significant differences (p=0.002). An overall variance of 6.2 (LLOA −2.9 to ULOA 15.4) was identified between reference images. Echogenicity accounted for the largest explained variance in muscle thickness (R^2^=0.81) and range of movement (R^2^=0.74), whilst muscle thickness and range of movement was the lowest (R^2^=0.61).

**Discussion:** People with FSHD demonstrated higher echogenicity, smaller muscle thicknesses and less range of movement. Histogram-matching for comparison of echogenicity values is necessary and can provide quantifiable differences. Different reference images affect echogenicity values but the variability is less than between group differences. Further work is needed to evaluate the longitudinal variability associated with this method on a larger sample of people with varying levels of arm function. Ultrasound scanning and post-histogram matching may be used to quantify and compare differences in muscle structure and function people with and without FSHD.

## Introduction

Facioscapulohumeral dystrophy (FSHD) is a common autosomal dominant muscular dystrophy with international prevalence estimates ranging from 0.8 to 12/100 000 ^1,2^. There are two genetic subtypes (FSHD1 and FSHD2), which have a common pathophysiological mechanism of ectopic expression of the DUX4 gene in skeletal muscle stemming from hypomethylation of the D4Z4 array ^3–5^. The natural history, presentation and progression speed of FSHD is highly variable between individuals which makes treatment planning complex. A common misconception, possibly reinforced by the disease name, is that it solely affects the muscles of the face, shoulders and arms, although this is not the lived experience of all people with FSHD ^6^. Some common phenotypes, based on patient reported symptoms, include impairments and disease trajectories considered ‘atypical’ for FSHD e.g. facial sparing variants or early onset of foot dorsiflexor weakness ^7^. Currently there are limited biomarkers which can be used clinically for subgrouping, prognosis planning, treatment evaluation and surveillance in FSHD.

Despite the heterogenous and asymmetric nature of FSHD, changes to the structure and function of the periscapular muscles is a hallmark feature for the majority of people affected by FSHD. Multiple signalling pathways and body systems are affected resulting in an inflammatory immune response with a cascade of oxidative stress, altered muscle cell differentiation in myogenesis and apoptosis ^5^. Body structures demonstrate oedema and notable degeneration of the muscle fibres, characterised by fatty and fibrotic muscle infiltration and increased fibre size variability ^8–10^. At the shoulder girdle, this results in a change to the biomechanical properties of the joint (sulcus), pain, progressive loss in range of movement, strength, function and independence ^6,11–13^. People with FSHD can also experience fatigue and respiratory complications which can negatively impact their overall functional capacity and upper-limb function ^5^.

Identification of biomarkers in the upper-limb, which are sensitive to disease progression, could be used for informing subgrouping and clinical decision making. Previous research has identified a relationship between structural changes (muscle echogenicity) observed in skeletal muscle during MRI that precede negative changes in function ^8^. However, MRI is not tolerated by all patients and has a considerable time and financial cost associated with it. Comparison between centres is also challenging in the absence of a reference phantom and variations in practice. Quantitative Muscle Ultrasound (QMUS) is being increasingly used as a biomarker in studies investigating FSHD ^8,14–17^. QMUS is a measure of echogenicity, used as an index outcome for inflammation and muscle structure changes (atrophy, fatty infiltration and fibrosis) which are relatively hyperechoic. QMUS has been shown to correlate with changes on MRI, FSHD clinical severity and qualitative ultrasound scores (Heckmatt scales) and precedes impairments and functional loss ^14–16,18,19^.

Despite the potential of QMUS, existing methods do not allow for clinically feasible comparisons as they are dependent on large reference datasets ^19,20^, consistent measurement settings within and across machines and are subject to ‘black box’ parameters of individual machines and models ^14,15,17^. Given the length of the disease course, lifespan of machines and rate of ongoing technological developments these methods may not be clinically feasible for assessing disease progression. Post image capture, it is possible to use mathematical methods for extracting or standardising different features of ultrasound images enabling comparison ^21^. Histogram-matching, is one such method which allows for the normalisation of images, as the histogram distribution of a target ultrasound image is transformed to match that of a reference image ^21^. Quantified echogenicity scores, measured using grayscale can then be measured across ultrasound images. Histogram-matching may therefore help overcome existing shortcomings of conventional QMUS ^20,21^. Evaluation of the periscapular muscles which are linked to upper-limb function using this method may be used to inform clinical decision-making regarding subgrouping, evaluation of treatments, prognosis planning and surveillance. However, a fundamental step in this process is evaluating the sensitivity of the measurements derived from this method and their relationship to other measures of body structure and function. Therefore, the aim of this study was to investigate if ultrasonography and post processing histogram-matching can be used to measure muscle echogenicity for differentiating between people with and without FSHD, and different levels of arm function. This paper reports the findings of ultrasound measurements for the trapezius muscle of people with FSHD and an age- and sex-matched control group as it was possible to extract information regarding muscle thickness and echogenicity for all participants.

## Methods

Ethical approval for this study was gained from the West Midlands - Black Country Research Ethics Committee 21/WM/0275.

### Study design

This was a single measurement, case control study. Participants were recruited from two separate sampling frames. These were a group of people with FSHD and an age- and sex-matched control group (CG). All participants were recruited from a single tertiary centre and through advertising across regional specialist centres. Recruitment was over a 12-month period. Five out of 19 people approached for the study declined or were unable to take part (recruitment rate 74%).

Participants who gave us informed consent attended a single measurement session during which demographic data, clinical measures, 2D-ultrasound imaging and 3D-movement analysis of their upper-limb was done. The main outcomes for this study were echogenicity scores, muscle thickness and range of movement (ROM) determined using the maximum thoracohumeral elevation angle.

### Inclusion criteria

People above the age of 18 were included for both groups. For the FSHD group, a confirmed diagnosis of FSHD was required. As this was a proof of concept study, no formal sample size calculation was conducted, however sample size was consistent with previous studies ^16,17,22^. Stratified sampling by arm function was used for subgrouping of: those able to lift their arm above shoulder height, those unable to lift arm above shoulder height and those with previous scapulothoracic arthrodesis. Controls were age- and sex-matched.

### Exclusion criteria

For the FSHD group, participants were excluded if there was any recent trauma to the shoulder within the last three months that had not resolved, surgery to the thorax or upper limb in the last six months, a previous history of fracture to the shoulder joint or any co-existing neurological pathologies or additional musculoskeletal injuries to the upper-limb being assessed. For the CG they were excluded if they had any previous presentation to a health care professional with a diagnosis of shoulder instability, a previous shoulder injury within the last three months that had not resolved, any co-existing neurological pathologies or deficits, any previous surgical interventions on the arm or were undergoing or awaiting medical management, diagnostic investigations on the arm.

### Demographics and clinical assessments

Patient demographics, Beighton scores of hypermobility and grip strength testing were recorded in addition to a clinical assessment of the shoulder (Appendix 1).

### 2D ultrasound measurement protocol

Ultrasound images of additional upper-limb muscles and structures were taken at the measurement session (Appendix 1) but only the trapezius muscle was selected for analysis as it was possible to extract muscle thickness and echogenicity for all participants. The trapezius muscle supports control of the scapula movement, which is important for upper-limb function ^23^. For muscle thickness measurements a gel stand-off layer was used to minimise tissue compression.

Surface bony landmarks were used for determining the point of muscle thickness measurements. Measurement of the middle trapezius were taken at the midpoint of a line between the 7^th^ cervical vertebrae and the acromioclavicular joint. A total of six measurements were taken (3-longitudinal and 3-transverse views) using an Esoate MyLab-Gamma device and linear probe (3-13 MHz). Depth and focus were adjusted for individual participants to ensure sufficient depth and landmark identification. Muscle thickness measurements were taken prior to the 3D movement analysis measurements.

Post capture muscle thickness measurements were carried out using ImageJ 1.53t. In cases where the width image edges were affected by artefact, stemming from a lack of probe contact whilst trying to measure the muscle but not apply compression, the sections of the image unaffected by artefact were used. Muscle thickness was determined by measuring from the most inferior aspect subcutaneous fat layer (demarcating the superior border of the trapezius muscle) to the most superior aspect of the fascial plane separating the trapezius from the underlying supraspinatus muscle (demarcating the inferior border of the trapezius muscle). For longitudinal views, the trapezius muscle was uniform across the image, allowing for the image midpoint to be used for calculating muscle thickness. To determine the midpoint a line, orientated to the muscle fibres was drawn across the length of the available image and was then bisected by another orthogonal line, which was used to measure the thickness of the muscle at this point (figure 1).

**Figure 1.**
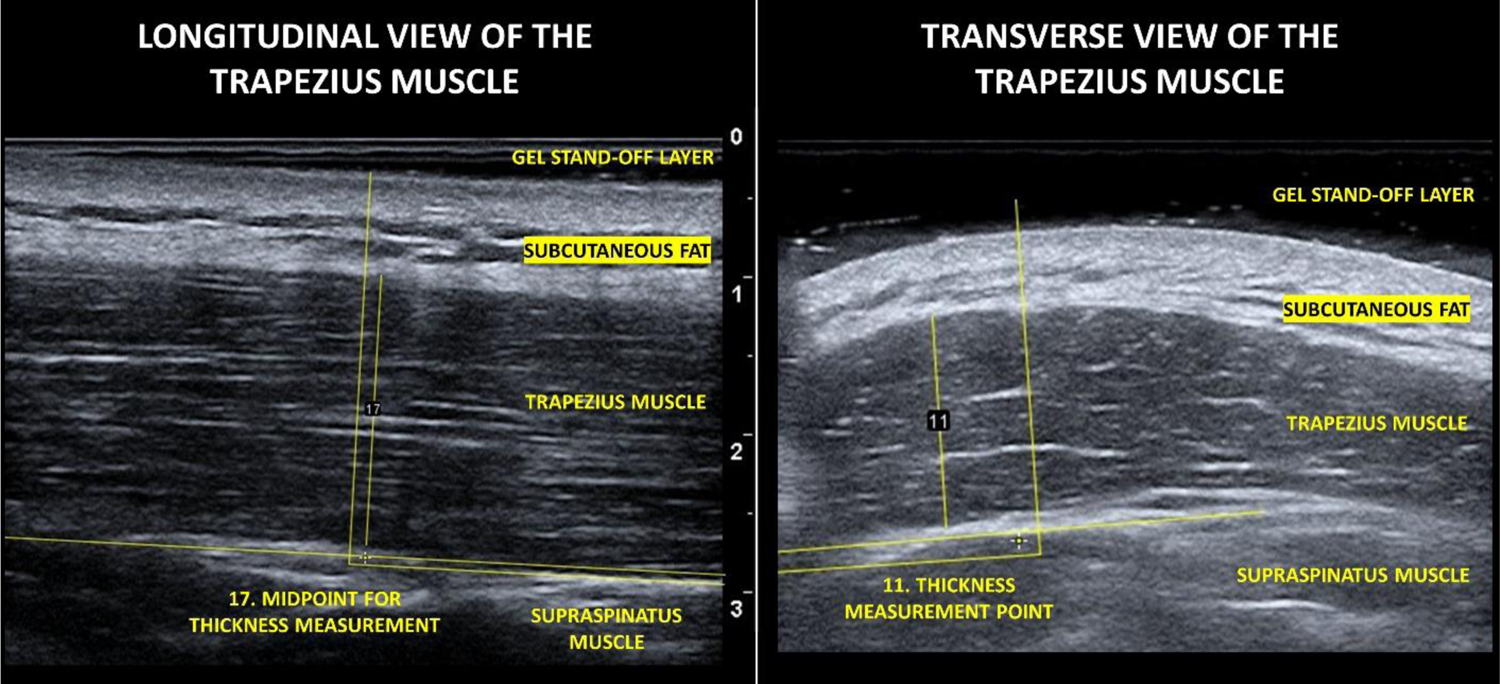
Methods for measuring trapezius muscle thickness in transverse and longitudinal views.

In the transverse view, the trapezius muscle was not uniform across the image for all participants (9 out of 15), therefore, the thickest part of the muscle was measured. For the transverse view, in six of the 14 participants, the midpoint was equivalent to the thickest part of the muscle. For measuring muscle thickness in the transverse view, a line orientated to the direction of the muscle fibres was drawn through the area of the image to be measured. An orthogonal line was then drawn at the thickest part to measure the thickness.

### 3D motion analysis protocol

Marker cluster, surface electromyography placement, static calibration processes, gap filling and filtering of kinematic and surface electromyography waveforms have been reported previously ^24^. Retroreflective marker clusters were placed on the thorax, acromion, humerus, forearm and hand segments ^25–27^ available at https://datacat.liverpool.ac.uk/2386/. Maximum thoracohumeral joint elevation angles are reported.

All movements were conducted in sitting and participants completed four unweighted upper-limb tasks (flexion, abduction, abduction to 45° with axial rotation, and hand to back of head) and three weighted tasks (self-selected) of 0.5kg, 1.0kg or 1.5kg (flexion, abduction, abduction to 45° with axial rotation) in that order. A total of 12 repetitions (2 sets x 6 repetitions) were carried out for unweighted tasks and a total of 6 repetitions (2 sets x 3 repetitions) were carried out for weighted tasks. Participants were initially shown the movements by the assessor and then asked to carry them out to a count of 3 seconds up, 3 seconds down, mirroring the assessor who was positioned in front of them.

Data were collected using a Vicon motion capture system at 100Hz (12 V5-Vantage motion analysis cameras, two synchronous coronal and sagittal video recordings and Delsys Trigno electromyography system sampling at 2000Hz).

### Data processing and analysis

Group demographic data are presented as frequencies. Histogram-matching was carried out as described by Bottenus et al. 2020 ^21^. All images were matched to a single reference image (transverse view of the trapezius muscle) from a CG participant, a younger male who undertook regular upper-limb physical activity. Manual segmentation of the subcutaneous fat layer was carried out and used as the region-of-interest for histogram-matching across all images (Appendix 2). Using full histogram-matching, the monotonic transformation was applied across the entire image. The trapezius muscle was segmented to determine echogenicity by quantifying mean grayscale values.

International Society of Biomechanics (ISB) recommendations were followed for joint co-ordinate system definitions and joint angles were calculated using inverse kinematics in Opensim 4.4 ^28–30^. Scaling ratios were determined from maker pairs associated with individual bony segments, identified in the static calibration and consistent with best practice guidelines ^24,31,32^.

### Sensitivity analysis

To evaluate the effect of histogram-matching on ultrasound images, echogenicity values of ultrasound images with and without normalisation were compared. To explore the sensitivity of the histogram matching method based on different reference images, analysis was carried out using a Bland-Altman plot^33^. The originally selected reference image (transverse view) was compared against a longitudinal view of the same muscle for the same participant, regions of interest and capture settings.

### Comparison of groups for echogenicity scores

A student t-test was used to determine if between group echogenicity values, (measured from ultrasound images normalised to the transverse reference image), were statistically significant with the alpha threshold set at 0.05.

### Relationship between muscle structure and function

The relationship between trapezius muscle thickness and echogenicity was explored by plotting the average muscle thickness across all views with the average echogenicity values across all views. The relationship between trapezius muscle structure (echogenicity) and function (ROM) was explored by plotting the average muscle thickness across all views against the maximum thoracohumeral angle of elevation from the abduction with a weight movement. Abduction was selected after Spearman’s correlation was used to identify the movement and plane with the highest correlation with echogenicity (Appendix 3). The relationship between trapezius muscle structure (thickness) and function (ROM) was explored by plotting the average muscle thickness across all views for and the maximum thoracohumeral angle of elevation achieved during the movement of abduction with a self-selected weight.

## Results

### Participant demographics

Data was collected for 14 participants, seven people with FSHD and seven age- and sex-matched controls. Demographic characteristics for all participants are reported in table 1. For the FSHD group, three people were able to lift their arm above shoulder height, two were unable to lift their arm above shoulder height and two had previous scapulothoracic arthrodesis.

**Table 1.** Demographic characteristics of study participants.

|  | Age matched controls (CG) (n=7) | People with FSHD (n=7) |
| --- | --- | --- |
| Age | 41.3 (15.5) | 41.9 (17.1) |
| Sex (M:F) | (5:2) | (5:2) |
| Height | 176.4 (5.7) | 176.0 (8.8) |
| Weight | 77.1 (11.2) | 90.6 (24.8) |
| Beighton score (median (IQR)) | 1 (0 – 2) | 0 (0 – 1.5) * |
| Grip strength Mean max value left (kgf) | 43.1 (7.9) | 31.1 (12.3) |
| Grip strength Mean max value right (kgf) | 48.1 (6.7) | 24.9 (11.8) |
| Dominant hand (L:R) | (1:6) | (2:4) |
| Weight selection for loaded tasks (0.5kg:1.0kg:1.5kg) | (0:0:7) | (1:4:2) |

\* 2 participants unable to complete Beighton due to standing balance issues

Results for maximum ROM across all activities at the thoracohumeral joint for the FSHD group and the CG are presented in table 2. The CG achieved larger ROM values when compared to the FSHD group. This is anticipated based on the use of stratified sampling by arm function.

**Table 2.**
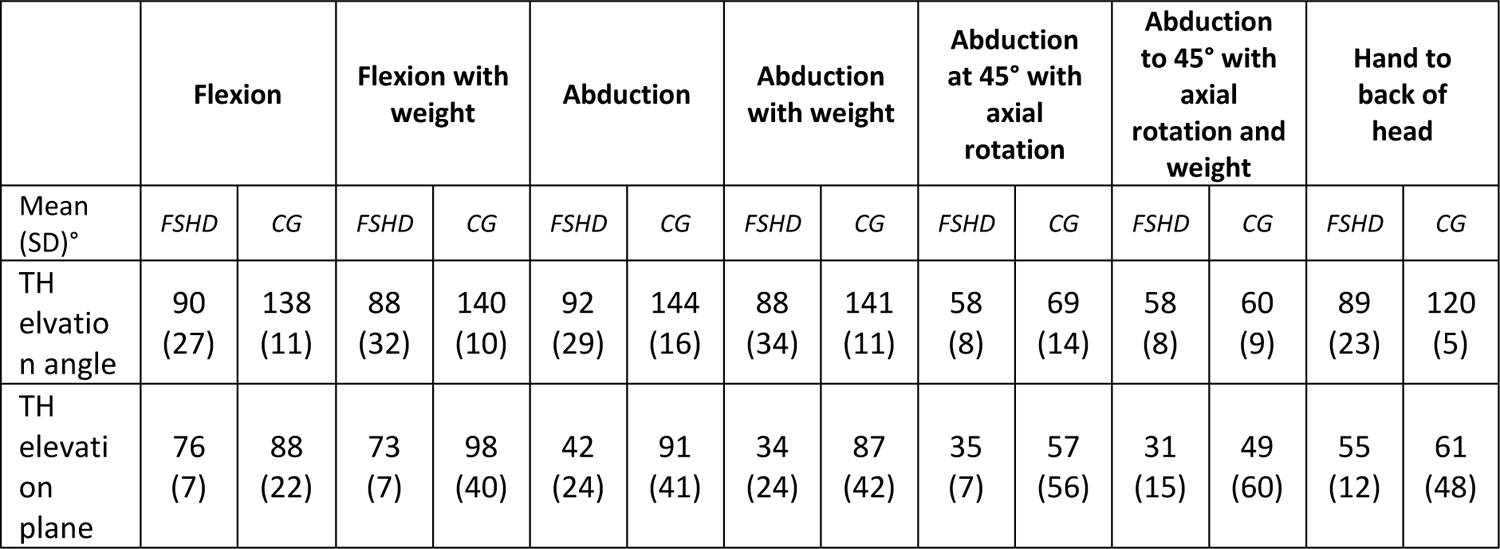
Maximum range of movement values for the thoracohumeral joint of pwFSHD and the CG.

|  | Flexion |  | Flexion with weight |  | Abduction |  | Abduction with weight |  | Abduction at 45° with axial rotation |  | Abduction to 45° with axial rotation and weight |  | Hand to back of head |  |
| --- | --- | --- | --- | --- | --- | --- | --- | --- | --- | --- | --- | --- | --- | --- |
| Mean (SD)° | FSHD | CG | FSHD | CG | FSHD | CG | FSHD | CG | FSHD | CG | FSHD | CG | FSHD | CG |
| TH elevation angle | 90 (27) | 138 (11) | 88 (32) | 140 (10) | 92 (29) | 144 (16) | 88 (34) | 141 (11) | 58 (8) | 69 (14) | 58 (8) | 60 (9) | 89 (23) | 120 (5) |
| TH elevation plane | 76 (7) | 88 (22) | 73 (7) | 98 (40) | 42 (24) | 91 (41) | 34 (24) | 87 (42) | 35 (7) | 57 (56) | 31 (15) | 49 (60) | 55 (12) | 61 (48) |

The plane of elevation was similar between groups for most of the movements apart from unweighted and weighted abduction. The FSHD group may likely be changing plane of elevation as a compensation method for achieving more ROM in abduction.

### Sensitivity analysis

A box and whisker plot of echogenicity scores derived from images with and without normalisation for both groups are presented in figure 2. Echogenicity scores derived from non-normalised images were generally higher and more variable resulting in a lesser ability to distinguish between groups (Figure 2). For the normalised images Mean (SD) echogenicity values for the FSHD group were higher than the CG (118.2 (34.0) vs 42.3 (14.0) respectively) with statistically significant differences observed (p=0.002).

**Figure 2.**
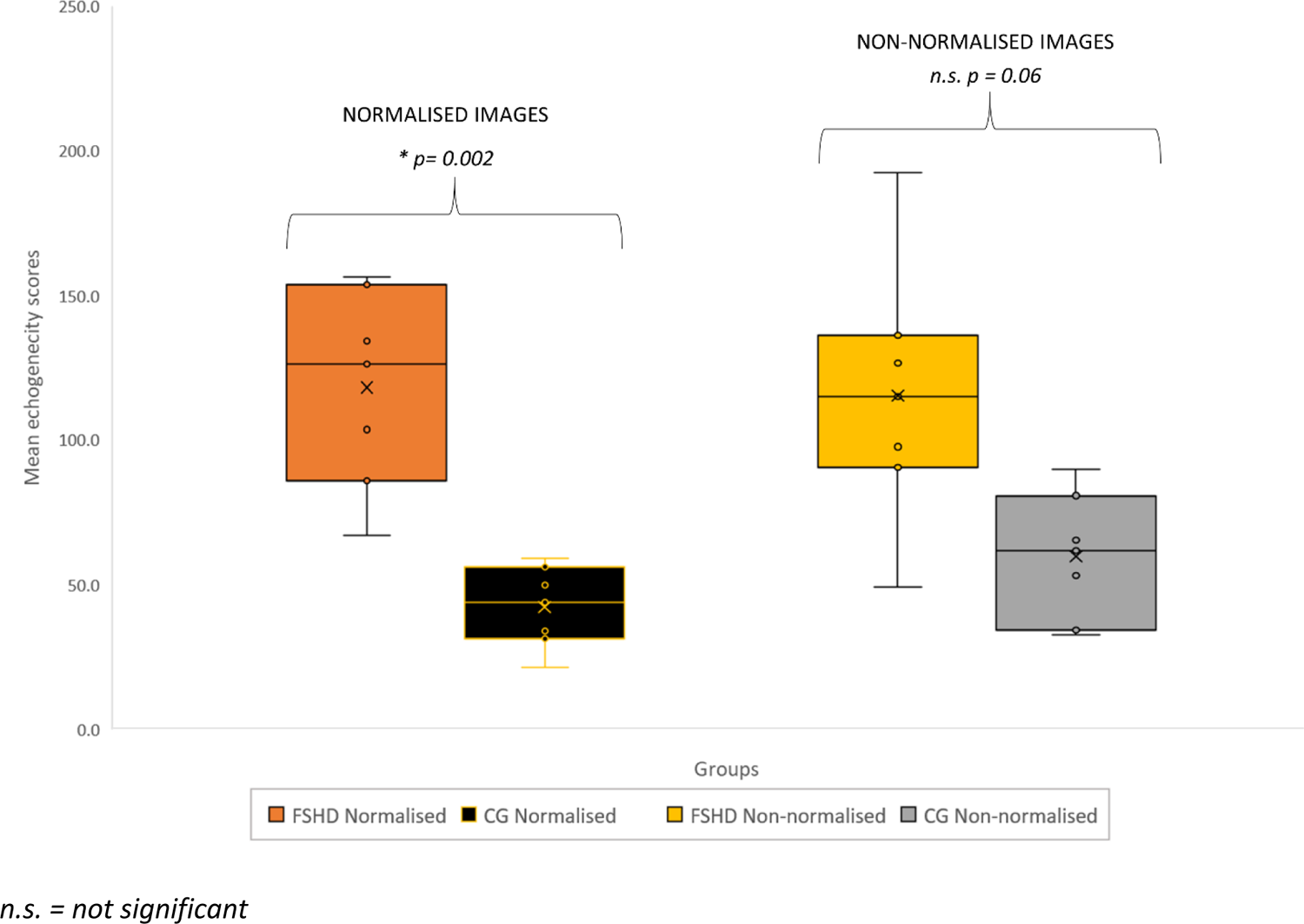
Mean echogenicity values for all groups, derived from both normalised and non-normalised ultrasound images.

Results for the sensitivity analysis echogenicity scores based on different reference images are presented in figure 3.

**Figure 3.**
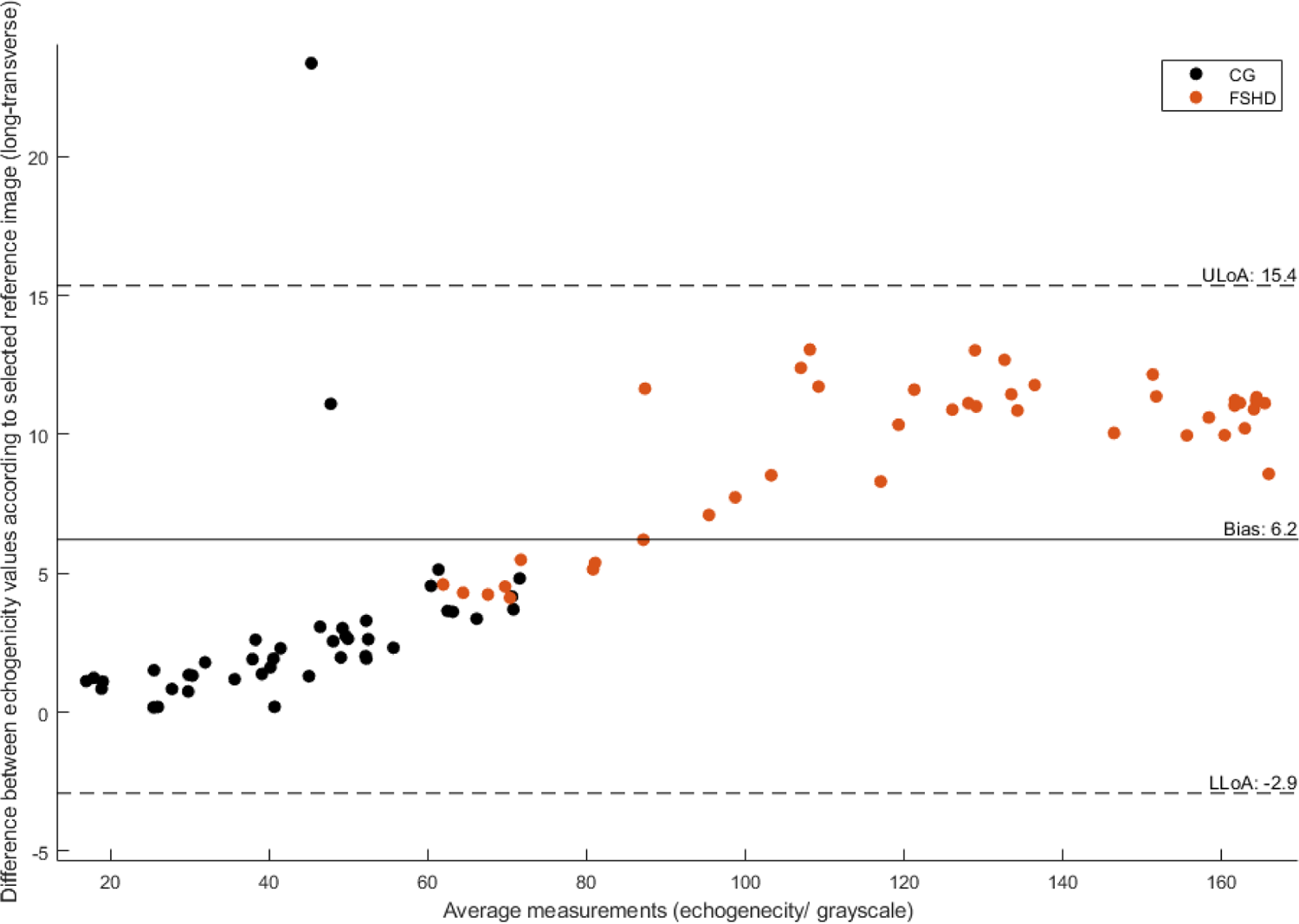
Bland-Altman Plot for trapezius muscle echogenicity scores derived using different reference images and histogram matching.

The Bland-Altman plot shows an overall variance of 6.2 between the longitudinal and transverse images for echogenicity scores (lower level of agreement −2.9 and upper level of 15.4). The longitudinal reference image was associated with a higher echogenicity score offset relative to the transverse image. The largest differences were observed for the FSHD group and people with FSHD who’s images overall had higher echogenicity scores. In cases where people with FSHD had similar levels of arm function to their age- and sex-matched control, difference between images were similar. Overall the variance between reference images were smaller for the CG.

### Relationship between muscle structure and function

Results outlining the relationship between different measures of upper-limb structure and function are presented in figure 4.

**Figure 4.**
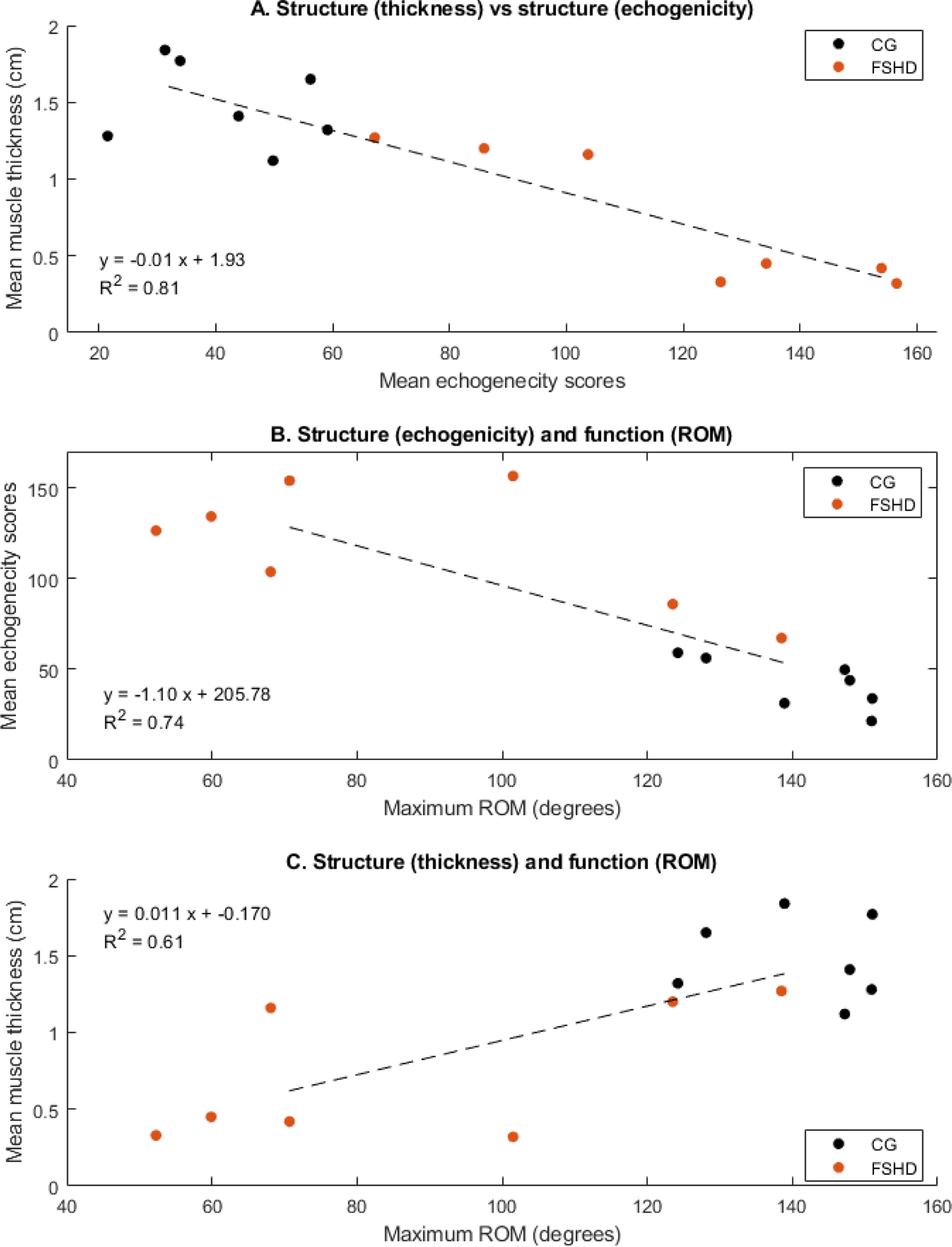
Relationship between muscle structure and function.

The largest explained variance was observed for muscle thickness and echogenicity (R^2^=0.81), followed by echogenicity and ROM (R^2^=0.74). Muscle thickness and ROM had the lowest explained variance (R^2^=0.61) possibly indicating that muscle thickness has limited capacity for explaining the variance in muscle function (maximum ROM). Results suggest echogenicity scores are better at accounting for the variance in muscle thickness values and maximum ROM. In most cases, a distinction between groups is evident based on the measurements evaluated, with mean (SD) muscle thickness values being higher in the CG compared to the FSHD group (1.48 cm (0.27) vs 0.74 cm (0.45)) respectively and the CG being able to achieve higher ROM for weighted abduction compared to the FSHD group (141° (11) versus 88° (34) respectively).

## Discussion

The aim of this study was to investigate if ultrasonography and post processing histogram matching can be used to measure muscle echogenicity as a biomarker for disease progression, for differentiating between people with and without FSHD, and different levels of arm function. Within our study, people with FSHD demonstrated higher echogenicity and smaller muscle thicknesses values indicative of degenerative muscle structure changes such as atrophy, fatty infiltration, fibrosis and oedema associated with the disease. Results suggest that post-capture processing of ultrasound images using histogram-matching is needed for comparison of echogenicity values and can provide quantifiable differences in people with and without FSHD. An estimation of the error based on the use of different reference images was also identified, which may be used for informing comparison or interpretation of images for decision making.

Evaluation of the trapezius muscle identified that the FSHD group had higher echogenicity values and less ROM. Our results were consistent with similar studies evaluating the relationship between echogenicity of the trapezius muscle and levels of impairment in people with FSHD ^14,15^. The mean (SD) values observed in our cohort of seven pwFSHD was 118.2 (34.0), which was higher, but similar than those of Goselink et al. 2020 who measured 22 symptomatic pwFSHD longitudinally (80% of which had affected trapezius muscles) and reported mean echogenicity values of 96.36 and 101.3 at baseline and 12-months respectively ^15^. In this case higher echogenicity values were associated with increased disease severity. Despite similarities between our findings and other research regarding echogenicity values, methods for measuring echogenecity varied. In our study histogram-matching, was used and image capture settings varied whilst in other studies, machine parameters had to remain constant ^14,15^. An advantage of our selected method is that it allows for measurement settings to be optimised for individuals and comparisons across models and machines may be possible. The requirement for consistent machine model type and normative reference data sets limits retrospective and cross study comparison, more widespread use and translation into clinical practice. Use of a standard reference image and post-processing, alongside an understanding of the margins of error could address these factors.

Echogenicity values are subject to variation dependant on the reference image used, however these were small (bias of 6.2) and less than the between group differences observed in our study. Differences/error between the reference images for calculating echogenicity scores were larger in the FSHD group. An offset for higher echogenicity values was identified when the longitudinal reference image was used. This offset possibly stemmed from the presence of more hyperechoic regions associated the muscle fibres potentially resulting in increased signal attenuation across the whole image and region of interest (subcutaneous fat layer) used for histogram matching. An assumption of our study was that the distribution of the subcutaneous fat layer was homogeneous within and across participants and this was supported by preliminary analysis of the methods used. If differences in the distribution was present within the subcutaneous fat layer region of interest, this may affect results between participants but not within the same participant. All muscle thickness measurements, manual segmentation of subcutaneous fat and muscle regions of interest were taken by a single assessor trained in ultrasound and image segmentation. Application of the post-histogram matching (dependant on the reference image) was consistent across all regions of interest and any variability likely stems from the reference image itself. Variability in image segmentation regions of interest for reference or target images may affect echogenicity values, particularly if conducted by another assessor. Further work is needed to investigate the inter-rater reliability for image segmentation and evaluation of echogenicity scores.

Upper-limb echogenicity values, derived from ultrasound and histogram-matching could facilitate clinically feasible bedside methods for assessing and monitoring disease progression in people with FSHD. Echogenicity has been used as a biomarker for screening and motoring in studies investigating FSHD, other neuromuscular diseases ^34,35^ and settings such as Intensive Care units ^36^. Our study identified differences in echogenicity scores between groups which were statistically significant. Whilst other methods such as the Heckmatt scale are available for classifying changes to muscle structure, and correlate well with QMUS, it an ordinal score which does not allow for quantification of echogenicity and is unable to determine changes to muscle structure within existing classification categories ^14,15^. Whilst QMUS may be a valuable biomarker in the assessment and management of people with FSHD, it is important to note that it is a local measure, limited by beam width and variable performance in some patient types i.e. those with higher adipose levels. Fatty infiltration in muscle structure for FSHD is heterogenous, with a potential proximal to distal progression as determined by MRI ^16^. Given that QMUS is often conducted on the mid muscle belly, not scanning the entire muscle structure may result in omission of some muscle structure changes ^14–16,37^. In some longitudinal studies, echogenicity scores in a limited number of pwFSHD have reduced (suggesting improvement to muscle structure), however, variations in echogenicity values may stem from fluctuations in levels of swelling as well as fibrosis which QMUS is unable to differentiate ^14^. Interpretation of QMUS measures therefore needs to be undertaken with the understanding that measurement location, method of analysis and fluctuations in immunochemical disease processes may affect echogenicity scores.

The trapezius muscle is one of the more commonly evaluated upper-limb muscles for FSHD and demonstrates changes consistent with disease progression ^14,15^. Given the role of the trapezius muscle in controlling the scapula and relative ease of measurement due to its size and morphology, compared to muscles such as serratus anterior, it may be a pragmatic choice for monitoring and prognosis planning in the upper-limb of people with FSHD ^23^. Within our study, the trapezius muscle was selected as it allowed for evaluation of muscle thickness and echogenicity for all participants. In cases where participants had smaller body segments or muscles, particularly for people with FSHD, capturing images that sufficiently showed the entire thickness and did not compress the muscle was challenging. This was because in some cases the requirements to maintain sufficient surface contact with linear probe and lesser convex surface of the underlying body segment resulted in loss of contact. Previous studies, including ours have used a limited subset of upper-limb muscles for evaluation of echogenicity scores and changes to function or disease progression. Within our study echogenicity was able to explain 74% of the variance in ROM (R^2^ value = 0.74) and further evaluation of other upper-limb muscles involved with control of the shoulder girdle and arm may help explain the outstanding variance, although determining consistent methods, including location of structures and positions for patients, can be challenging to standardise. Sites for measurement, derived with reference to surface bony landmark were selected in our study as a pragmatic method for standardising measurements rather than use of respective anatomical reference points within a muscle measured by ultrasound as these may be subject to operator variability. Furthermore, whilst several studies have evaluated the trapezius muscle, it is possible that the site of measurement within the muscle may vary between studies and methods for determining the measurement point were not explicit in all studies ^14,15,17^. Future work may look to agree standardised protocols and muscles for QMUS that could be reproduced and possibly used to inform screening, prognosis, monitoring and clinical decision making.

Measurement of both muscle echogenicity and thickness values may be required for monitoring and prognosis planning in the upper-limbs of people with FSHD. QMUS was able to account for a large proportion of the variance in muscle thickness (structure) and maximum ROM (function) with R^2^ values of 0.81 and 0.74 respectively. These findings may be anticipated given that 1) changes in one structural component of a muscle (e.g. fibrosis measured by echogenicity) will likely be reflected in another (e.g. thickness) 2) trapezius is known to be a significant contributor for ROM at the joint 3) redundancy, stemming from the number and thickness of upper-limb muscles may account for the limited relationship between measures ^38^. Whilst muscle thickness was able to account for some of the variance in ROM R^2^ = 0.61, this was limited when compared to other measures. Additional measures of force production, which is proportional to muscle thickness may increase the explained variability e.g. Dijkstra et al. 2021 found a correlation between echogenecity and strength in children with FSHD of r = −0.74 ^14^. Previous research has reported variable levels of correlation (approximately 0.5 to 0.6) with QMUS and the FSHD clinical severity scale ^14,15^, however lower levels of agreement may be attributed to the limited measurement properties of the FSHD clinical severity ordinal score rather than QMUS. Use of muscle thickness measures in isolation for prognosis planning or categorisation of function may therefore be of limited value.

## Limitations

This was a single measurement study and has not evaluated the longitudinal variability or changes of measures or methods used. Further work is needed to evaluate the longitudinal variability associated with this method on a larger sample of people with varying levels of arm function. This should also include variability in the echogenicity scores based on multiple images taken from different sites within a muscle and with analysis of image segmentation and histogram matching carried out by multiple assessors. Whilst theoretically the histogram-matching methods should allow for comparison between machine types, models and different muscles within and between people further work should evaluate this in order to facilitate a better understanding of differences which could be used to inform decision making. We were unable to compare our images against another imaging modality such as MRI which is considered the gold standard ^15^. QMUS has already been evaluated against MRI, using different echogenicity analysis methodology and shows high levels of agreement, despite a lesser ability to differentiate swelling and fibrosis within ^16,18^. Echogenicity scores are variable subject to the size and region selected. Whilst segmentation was possible for all participants, this was challenging in cases where participants either had very small subcutaneous fat layers (mainly CG), small muscles or edge artefact present in some components of the muscle. Whilst segmentation was carried out to include only relevant tissues with no artefact and were manually checked for spurious results possibly indicating errors, it is acknowledged that these potentially could have affected results for individual people. The same is true for muscle thickness measurements. A comparison of two reference images was carried out and it is unclear how the magnitude of the error varies across multiple images of the same or different muscle. The reference image selected was chosen on the basis that it came from one of the younger CG participants who engaged in regular upper-limb exercise and would therefore be a suitable comparison for people who were older or had less function. Further work may look to compare multiple reference images and the effect on echogenicity scores, possibly in multiple neuromuscular diseases. Whilst QMUS and the use of post-histogram matching has demonstrated an ability to differentiate between groups with and without FSHD it should not be considered as a diagnostic tool given that the accuracy of this method has not been evaluated in real-world clinical settings.

## Conclusion

Histogram-matching for normalisation of ultrasound imaged are better able to differentiate and quantify difference between people with and without FSHD than non-normalised images. The between group differences observed were larger than the error of measurement calculated using two reference images. Overall echogenicity scores derived from the trapezius muscle, were able to better account for the variance in muscle thickness values and function. Muscle thickness had limited ability in accounting for function when evaluated in isolation. The FSHD group had higher echogenicity values and smaller muscle thicknesses when compared to the CG, indicative of degenerative muscle structure changes associated with the disease. Ultrasound of the trapezius, and selection of measurement site based on surface bony landmarks allowed for pragmatic simultaneous capture of measurement and thickness in all participants. This could facilitate clinically feasible bedside methods for assessing and monitoring disease progression in people with. Further work is needed to recruit a larger sample of people with FSHD and varying levels of arm function, carry out longitudinal measurements and evaluate the sensitivity of these measures on the basis of variable reference images.

## Data Availability

All data produced in the present study are available upon reasonable request to the authors

https://datacat.liverpool.ac.uk/

