## Supplementary figures and images for "Quantifying morphological changes in middle trapezius with ultrasound scanning and a novel histogram-matching algorithm for adults with and without Facioscapulohumeral dystrophy (FSHD)"

### Appendix 1

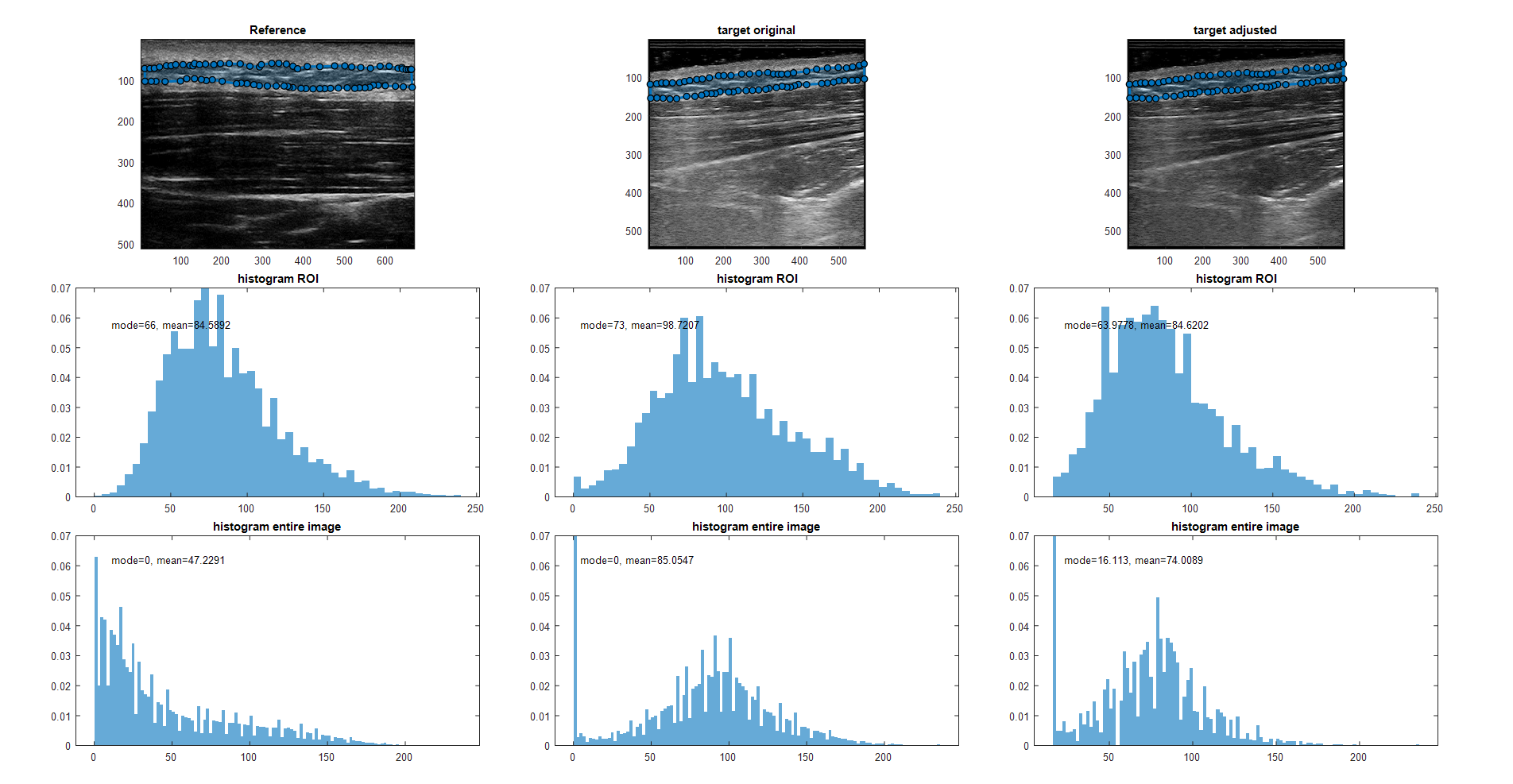

### Appendix 2

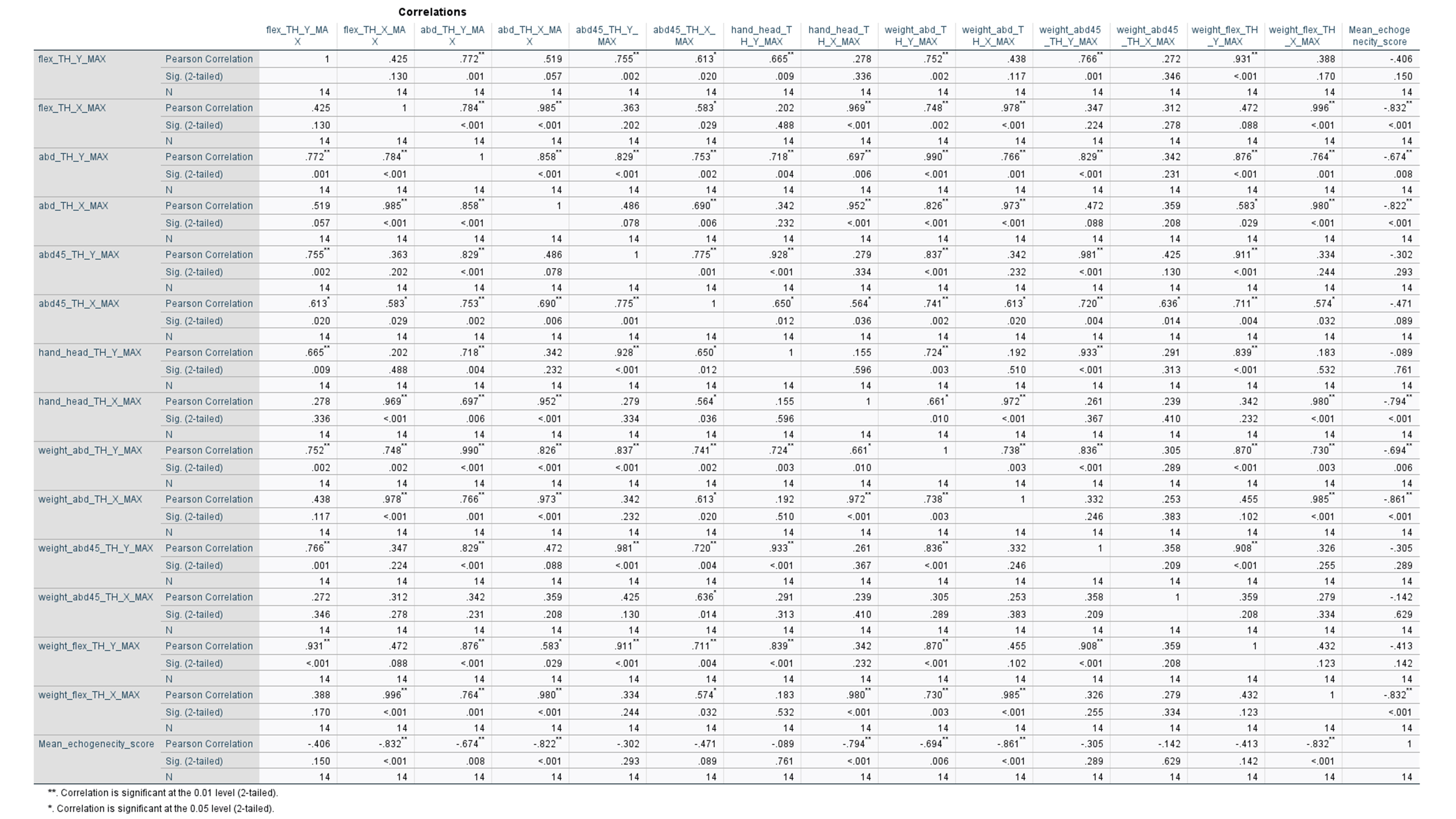
