## Appendix 3 for "Quantifying morphological changes in middle trapezius with ultrasound scanning and a novel histogram-matching algorithm for adults with and without Facioscapulohumeral dystrophy (FSHD)"

**Appendix 1. Muscles and structures evaluated during measurement session.**

**Demographics and clinical assessments**

Clinical assessments included recording of the following clinical features of the shoulder: apprehension, guarding or laxity in the sulcus, anterior and posterior shift load, and apprehension relocation tests, Beighton scores of hypermobility and grip strength testing.

**2D ultrasound measurement protocol**

Ultrasound of the upper-limb was carried out to explore the feasibility of measuring different upper-limb muscles and anatomical structures in people with FSHD. Muscle images included three longitudinal and three transverse views of the

- Trapezius
- Supraspinatus
- Infraspinatus
- Biceps brachii
- Triceps and
- Anterior deltoid muscles.

Anatomical structure images included the

- Long head of biceps
- Pectoralis major tendon
- Subscapularis tendon
- Supraspinatus
- Subacromial space and
- Posterior glenohumeral joint recesses.
